# Refinement of a published gene-physical activity interaction impacting HDL-cholesterol: role of sex and lipoprotein subfractions

**DOI:** 10.1101/2024.01.23.24301689

**Authors:** Kenneth E. Westerman, Tuomas O. Kilpeläinen, Magdalena Sevilla-Gonzalez, Margery A. Connelly, Alexis C. Wood, Michael Y. Tsai, Kent D. Taylor, Stephen S. Rich, Jerome I. Rotter, James D. Otvos, Amy R. Bentley, Samia Mora, Hugues Aschard, DC Rao, Charles Gu, Daniel I. Chasman, Alisa K. Manning, the CHARGE Gene-Lifestyle Interactions Working Group

## Abstract

Large-scale gene-environment interaction (GxE) discovery efforts often involve compromises in the definition of outcomes and choice of covariates for the sake of data harmonization and statistical power. Consequently, refinement of exposures, covariates, outcomes, and population subsets may be helpful to establish often-elusive replication and evaluate potential clinical utility. Here, we used additional datasets, an expanded set of statistical models, and interrogation of lipoprotein metabolism via nuclear magnetic resonance (NMR)-based lipoprotein subfractions to refine a previously discovered GxE modifying the relationship between physical activity (PA) and HDL-cholesterol (HDL-C). This GxE was originally identified by Kilpeläinen et al., with the strongest cohort-specific signal coming from the Women’s Genome Health Study (WGHS). We thus explored this GxE further in the WGHS (N = 23,294), with follow-up in the UK Biobank (UKB; N = 281,380), and the Multi-Ethnic Study of Atherosclerosis (MESA; N = 4,587). Self-reported PA (MET-hrs/wk), genotypes at rs295849 (nearest gene: *LHX1*), and NMR metabolomics data were available in all three cohorts. As originally reported, minor allele carriers of rs295849 in WGHS had a stronger positive association between PA and HDL-C (*p_int_* = 0.002). When testing a range of NMR metabolites (primarily lipoprotein and lipid subfractions) to refine the HDL-C outcome, we found a stronger interaction effect on medium-sized HDL particle concentrations (M-HDL-P; *p_int_*= 1.0×10^-4^) than HDL-C. Meta-regression revealed a systematically larger interaction effect in cohorts from the original meta-analysis with a greater fraction of women (*p* = 0.018). In the UKB, GxE effects were stronger both in women and using M-HDL-P as the outcome. In MESA, the primary interaction for HDL-C showed nominal significance (*p_int_* = 0.013), but without clear differences by sex and with a greater magnitude using large, rather than medium, HDL-P as an outcome. Towards reconciling these observations, further exploration leveraging NMR platform-specific HDL subfraction diameter annotations revealed modest agreement across all cohorts in the interaction affecting medium-to-large particles. Taken together, our work provides additional insights into a specific known gene- PA interaction while illustrating the importance of phenotype and model refinement towards understanding and replicating GxEs.

## Background

Gene-environment interactions (GxEs), which describe the modification of environmental effects on a phenotype by a genetic factor (or vice versa), inform attempts to better understand complex and polygenic traits such as cardiometabolic diseases and their risk factors. The emergence of larger-scale cohorts and consortia have allowed for hypothesis-free GxE discovery efforts in genome-wide interaction studies (GWIS). Such investigations have explored genetic modification of relationships such as that between fish oil supplementation and plasma lipids in the UK Biobank (UKB) cohort (Francis et al., 2021) or between and smoking and lipids in a large-scale meta-analysis (Bentley et al., 2019).

Despite increasing attention to GWIS, robust discovery and translation of their results faces multiple challenges. First, loci uncovered using such hypothesis-free approaches often lack evidence of clear biological mechanisms by which the genetic and environmental pathways overlap. This is an extension of the more general “variant to function” challenge in the genetics community, but with the added obstacle of incorporating an understanding of the interacting exposure. Second, replication of GxEs has proven to be a major challenge. Part of this replication challenge can be attributed to low statistical power for identifying interactions (Gauderman et al., 2017), which are a limiting factor for typical sample sizes used to-date (Westerman et al., 2023). Part is also due to heterogeneity across cohorts in the distribution and measurement of exposures as well as the complex confounding structure found in the observational datasets that are used for most GxE studies.

Both of these challenges can be addressed by “refinement” of GxEs originally identified in large-scale, hypothesis-free discovery efforts. A more fine-grained understanding of the specific interacting elements (e.g., low-versus high-intensity physical activity [PA]) and relevant sub-cohorts (e.g., GxEs that are specific to a given sex or ancestry subgroup) can inform more effective replication efforts, including cohort selection and modeling choices. Furthermore, omics measurements, such as metabolomics, have been used to interrogate mechanisms for marginal (interaction-free) genetic effects on complex phenotypes (Auwerx et al., 2023; Yin et al., 2022) and can further act as mediators of the genetic and/or exposure effects for an established GxE.

To explore the value of GxE refinement, we focused on the strongest GxE identified in a GWIS meta-analysis of PA on HDL-C (Kilpeläinen et al., 2019). We aimed to refine our understanding of its specific context using a series of observational datasets, structured modeling with additional covariates, and additional relevant data types (NMR-based lipoprotein measurements). In the Women’s Genome Health Study (WGHS) dataset, for which the interaction was strongest in the original meta-analysis, we first validated the interaction and identified the most relevant lipoprotein-related quantities affected. In the UKB dataset, we next explored the specificity of the interaction based on sex and physical activity subtypes. Finally, in the Multi-Ethnic Study of Atherosclerosis (MESA) dataset, we replicated the interaction while illustrating the potential for residual heterogeneity in comparing lipoprotein quantities across cohorts and measurement platforms.

## Results

### Outcome and population refinement in WGHS

WGHS was chosen to begin the investigation since it has (1) the most significant study-specific G×PA interactions at rs295849 in the original meta-analysis, and (2) available NMR metabolomics data. Data from 23,018 women were available with appropriate variables for analysis (population summary in Supp. Table S1). PA was associated with HDL-C in main effect (genotype-free) models (4% higher in active versus inactive, *p* = 1.5×10^-23^). The previously-reported interaction between rs295849 and PA (Fig. 1a) was not materially changed by the more substantial covariate adjustment used here. Sensitivity models with genetic principal components and with additional genetic interaction terms for each covariate showed no meaningful change in the primary interaction estimate (Fig. 1b).

**Figure 1:**
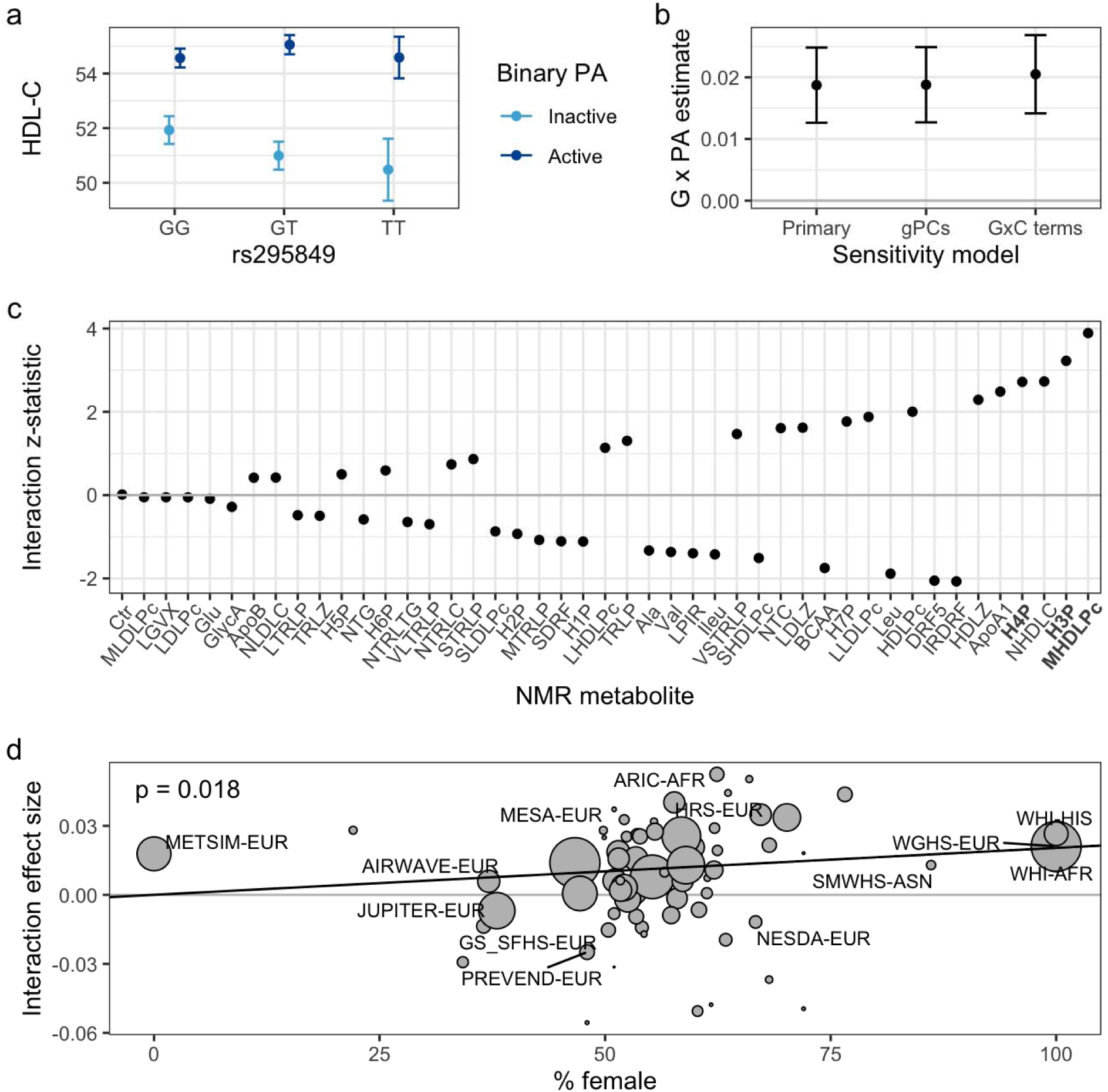
Validation and expansion of the known interaction in WGHS. a) Genotype- and PA- stratified HDL-C means (in mg/DL) show the interaction. b) Stability of the interaction in the context of additional covariates, with interaction effect estimates shown across multiple adjustment strategies (primary covariates, including genetic principal components, and including gene-covariate interaction terms for all covariates). c) Z-statistic of the primary interaction test using a series of alternative NMR-based outcomes (*x-*axis labels for M-HDL-P and its sub-components are bolded). NMR-measured HDL-C is indicated by “NHDLC” and a full list of metabolite abbreviations is available in Supp. Table S2. d) Bubble plot of study- and population- specific interaction estimates (in units of log-transformed HDL-C [mg/dL] / activity status / allele) contributing to the meta-regression. Interaction effect estimates are plotted against cohort sex proportions (percentage of females), with bubble sizes corresponding to interaction estimate precision (inverse of the effect estimate variance). Study labels are shown for studies with N > 2,000.

NMR-derived lipoprotein and other metabolites measures capture lipoprotein metabolism in greater detail than the clinical HDL-C measure alone. Thus, as alternative outcomes in otherwise identical statistical models, they can allow better resolution in understanding the relevant biology of the GxPA interaction. As shown in Fig. 1c and listed in Supp. Table S2, GxE significance was high for HDL-C but surpassed by medium HDL particle concentrations (M-HDL-P) and H3P (a subset of M-HDL-P).

The fact that the WGHS cohort is entirely female, and that HDL-C levels exhibit dimorphism according to sex, raises the possibility that the interaction may be more pronounced (i.e., a greater difference in PA effect across genotypes) in females compared to males. We performed a meta-regression on sex proportion to test this hypothesis, finding evidence that the interaction effect tended to be stronger in cohorts with a larger fraction of women (*p* = 0.018; I^2^ = 1.68%; Fig. 1d). This association was not due solely to the WGHS; in a sensitivity analysis excluding all single-sex cohorts, which would have outsized leverage affecting the meta-regression fit, the relationship became stronger (*p* = 0.0007; Supp. Fig. S1). Leave-one-out analysis confirmed that no single cohort was responsible for the effect, with the only substantial change being an increase in the meta-regression estimate and significance after removal of the all-male METSIM study.

### Exposure refinement and validation in UKB

A summary of the UKB population (N = 281,380 after exclusions and based on relevant data availability) can be found in Supp. Table S3. We first explored PA main effects in models without genotype terms. We verified the expected positive association between PA and HDL-C in the European-ancestry subset of UKB (0.10 SD HDL-C / SD PA, *p* < 10^-300^), which was present regardless of the questionnaire subsets used for PA estimation (IPAQ versus RPAQ; between-instrument Pearson correlation of 0.45; see Methods). Nested covariate adjustment sets showed that covariate choice affected the magnitude of the estimates: IPAQ-based PA estimates increased with more adjustment for SES and healthy lifestyle variables (possibly due to its inclusion of occupational PA, which often associates negatively with SES), whereas RPAQ- based measures decreased with more adjustment (indicating removal of the confounding effects of high SES and healthy lifestyle). For both questionnaire-based estimates, we observed a non-monotonic relationship in which HDL-C was generally positively associated with PA, but began to decrease at more extreme PA values (Supp. Fig. S2). We compared various PA transformations to account for this nonlinearity, using PA-HDL-C main effect strength as a guide, and chose to winsorize at the 90^th^ percentile for all PA variables moving forward (in both UKB and MESA). PA main effects were modestly different by sex (0.09 versus 0.13 SD HDL-C / SD PA in women and men, respectively; *p*-interaction = 7.8×10^-5^).

UKB provided an opportunity to explore the replication of the observed interaction across a series of modeling choices: NMR metabolite outcomes (as tested in the WGHS), female-only versus the full population (given the meta-regression results), and a more fine-grained set of potential PA variables (based on IPAQ and RPAQ). Interaction effects from these models are presented in Fig. 2a. In general, IPAQ-based PA estimates produced stronger effect estimates than those from RPAQ (mirroring results from PA-HDL-C main effect models; Supp. Fig. S2d), with no major differences evident using IPAQ subsets corresponding to moderate versus vigorous activity. Focusing on the IPAQ estimates, interaction effects on HDL-C (as measured by a standard biochemical assay) were somewhat greater in the subset of the population with available NMR data than the entire population. We did not identify any characteristics of the sub-population that clearly explained this heterogeneity (among those covariates listed in Supp. Table S3). As expected, effects estimated from NMR-measured HDL-C were comparable to those from standard HDL-C in the same sub-population (given the Pearson correlation of 0.94 between the two HDL-C measures). Finally, using the M-HDL-P outcome, we found that interaction effect estimates were modestly higher than for HDL-C and biased toward greater magnitudes in women, validating the outcome and sub-population refinements suggested by the WGHS analysis. Results for all NMR quantities are shown in Supp. Table S4. Visualization of the interaction using stratified genotypes and PA tertiles revealed a qualitative interaction mirroring that from WGHS, such that additional minor alleles amplified the positive PA-HDL relationship without inducing a meaningful marginal genotype effect (Fig. 2c). We note that differences in HDL subfraction labeling across NMR platforms complicates conclusions about concordance across cohorts (further discussion below).

**Figure 2:**
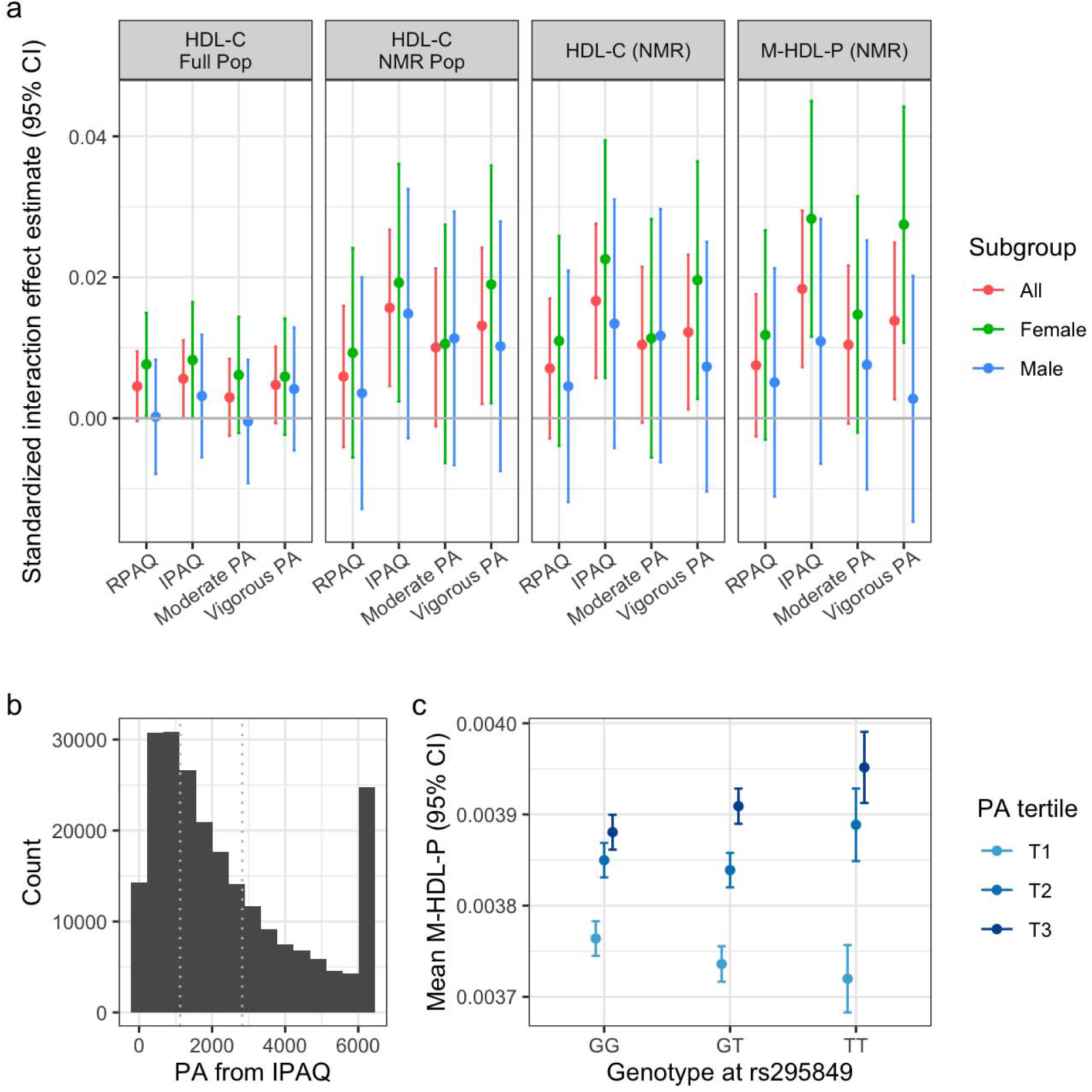
Refinement of the interaction in the UK Biobank. a) Interaction effect sizes (standardized to units of [SD outcome / SD PA / allele]) are plotted against various PA questionnaires and subtypes. Panels correspond to various outcomes and UKB sub-populations (full set of unrelated participants versus the subset with available NMR metabolomics data). b) Histogram of IPAQ-based PA measures, with the spike at the right reflecting the 90^th^ percentile winsorization. Dotted lines indicate tertile boundaries. c) Visualization of the interaction using mean M-HDL-P estimates plotted against allele at rs295849, stratified by tertile of IPAQ-based PA.

### Replication in MESA

We used the MESA cohort (N = 4,587) to test for replication of the interaction, including the refinements established in WGHS and UKB, noting that interaction effects in MESA population subgroups were directionally consistent but nonsignificant in the original meta-analysis. A summary of the MESA population can be found in Supp. Table S5. PA and HDL-C were positively associated (0.03 SD log(HDL-C) / SD PA, *p* = 0.01; Supp. Fig. S3), though less strongly than in the other cohorts, with no evidence of heterogeneity by sex. Despite a substantially smaller sample size than UKB and WGHS, limiting the statistical power for replication, MESA interaction models using NMR-measured outcomes showed nominally significant replication of the primary interaction influencing HDL-C (*p* = 0.01), but little indication of a female bias (Fig. 3a). Despite a consistent effect direction, the interaction did not affect M-HDL-P at a nominal significance level, rather showing an increasing interaction magnitude moving from small to medium to large HDL-P subfractions. After testing more granular HDL subfractions (seven categories based on average particle diameter), it appeared that the overall signal was attributable primarily to the largest particle subsets (H5P – H7P). Sensitivity analyses confirmed minimal heterogeneity in interaction effects when using PA subtypes (Fig. 3b) and when stratifying by self-reported race/ethnicity (Fig. 3c).

**Figure 3:**
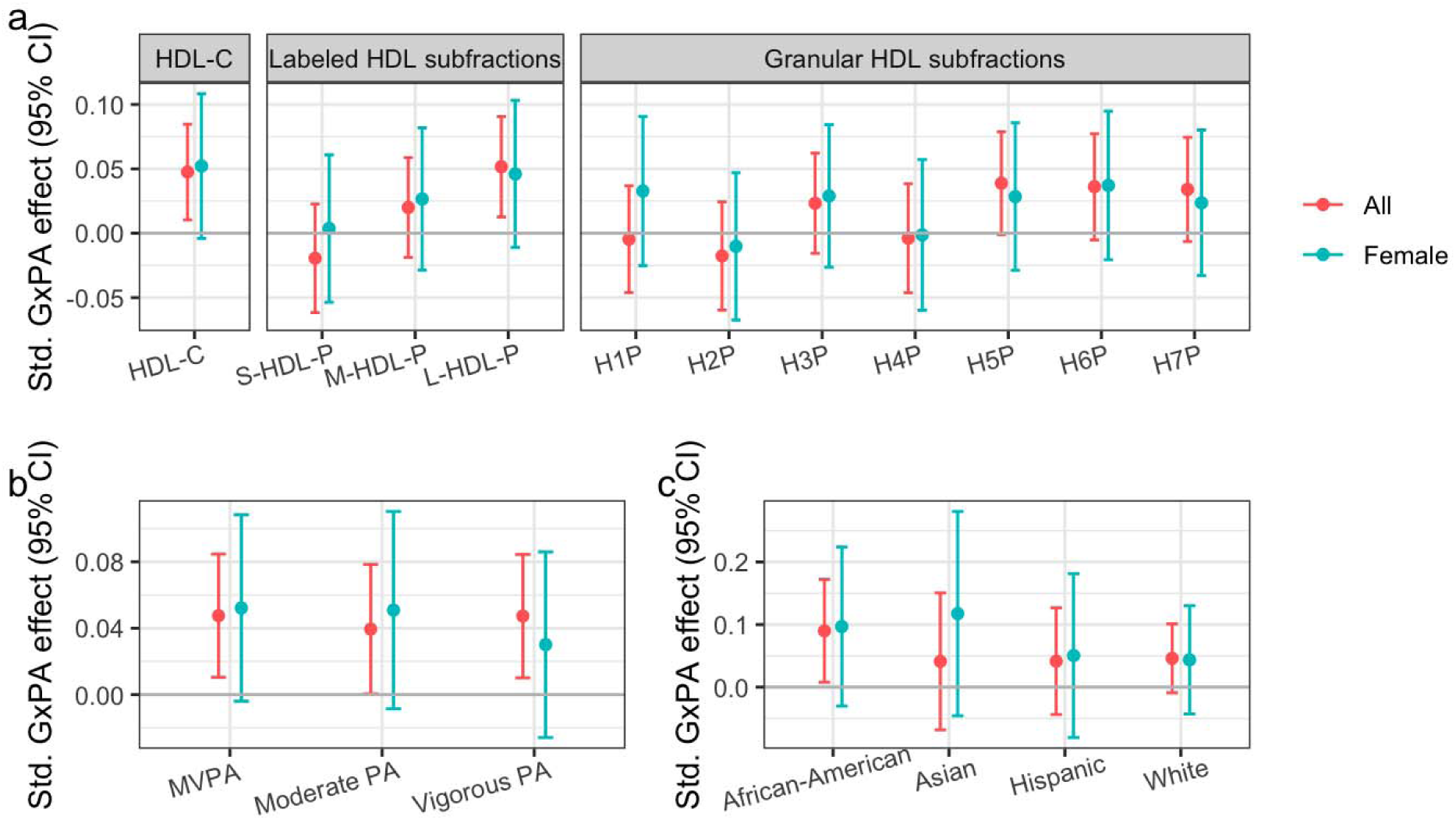
MESA replication results. a) Interaction effect sizes (standardized to units of [SD outcome / SD MVPA / allele]) are plotted against NMR-measured quantities including HDL-C, labeled HDL subfractions (small, medium, and large), and more granular HDL subfractions (in order of increasing diameter). b) As in (a), but plotting effects on HDL-C across PA types. b) As in (a), but plotting effects on HDL-C across PA types. c) As in (a), but plotting interactions involving MVPA and HDL-C across the four race/ethnicity subgroups comprising the MESA population.

As described above, we observed heterogeneity across cohorts in terms of the specific HDL-P subfractions showing the strongest signal. Given the difference in NMR measurement platform between UKB (Nightingale) and the others (Labcorp), we further explored the relationships between these subfractions and in the context of their specific particle size annotations. In MESA, larger HDL particles showed lower concentrations but higher correlations with HDL-C compared to smaller HDL particles, as expected (Fig. 4a). Importantly, we note major differences in subfraction labeling across platforms, such that the M-HDL-P subfraction from Nightingale platform corresponds more closely to the large, rather than medium, HDL-P subfraction reported by Labcorp (Fig. 4b). Integrating these objective particle size-related findings with our modeling results, we found that there was in fact closer agreement as to the most relevant particle size between UKB and MESA (around 11nm diameter particles), compared to WGHS (around 9nm). Integrating the results from all three cohorts, the interaction appeared to be more pronounced for a general group of medium-to-large HDL particles, with diameters between roughly 9 and 12nm.

**Figure 4:**
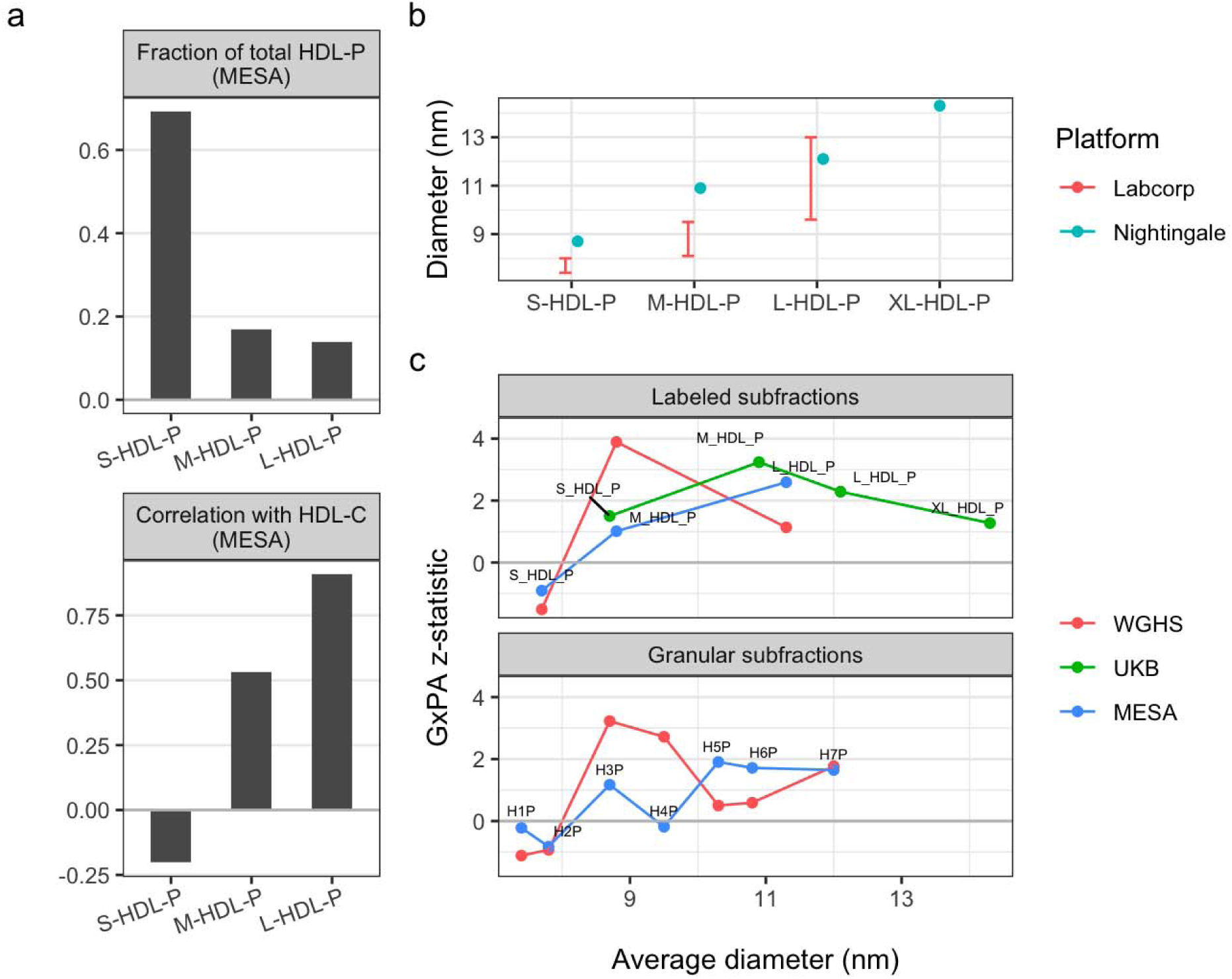
Exploration of HDL subfraction labels and sizes across platforms. a) Empirical relationships between Labcorp-labeled HDL subfractions in MESA, including fractions of overall HDL-P concentration (top panel) and Pearson correlations with NMR-measured HDL-C (bottom panel). b) Platform-specific particle diameters associated with HDL subfraction labels. Size-based annotations are available as ranges for Labcorp and as average diameters for Nightingale. c) Interaction testing z-statistics are shown as a function of annotated HDL size (rather than label) for each primary subfraction (small, medium, and large; top panel) and granular subfraction (H1P-H7P as reported by Labcorp only; bottom panel). Because WGHS and MESA were both measured using the Labcorp platform, labels are shown for MESA only. Average diameters for labeled Labcorp subfractions were approximated as the midpoint of the provided diameter range.

## Discussion

Our study was motivated by the recognition that even significant findings in large-scale, multi-study meta-analyses of GxE effects may provide only limited insight into mechanism. We chose one specific gene-PA interaction to explore, finding that increased attention to each piece of the interaction resulted in overall greater understanding and agreement among studies. Using WGHS and other cohorts from the original meta-analysis, we found that the interaction was stronger in women and impacted medium-sized HDL subfractions to a greater degree than HDL-C, conclusions that were supported by results in the much larger UKB study. MESA results replicated the primary interaction (with greater significance than its results in the original meta-analysis) and reinforced the tendency of the interaction to impact larger HDL particles, although without indication of a stronger effect in females as in the other cohorts.

Overall, we saw consistency in the impact of this interaction on HDL subgroups, with medium-to-large subfractions most strongly impacted, but there was some heterogeneity in the specific particle diameters affected. This is likely due to some combination of assay heterogeneity and biological variability between cohorts. Though both WGHS and MESA NMR data came from the Labcorp LP4 platform, effects were biased toward medium-sized particles (diameters between roughly 8-10 nm) in WGHS, versus larger particles (diameters greater than 10 nm) in MESA. UKB measurements came from the Nightingale platform, which uses a different set of diameter cutoffs and includes an “extra-large” subfraction. UKB results showed greater impacts on a diameter subset consistent with the L-HDL-P subfraction from MESA (average diameter of 10.9 nm). Taken together, these results indicate effects most strongly concentrated on medium-to-large HDL particles (approximately 9-11 nm in diameter), but with meaningful heterogeneity in this estimate across cohorts.

Medium and large HDL-P (as defined for the Labcorp panel, including relatively lower-diameter species) are more strongly associated with cardiovascular protection than smaller subfractions. M-HDL-P was the strongest predictor of reduced coronary events in statin-treated subjects from the MRC/BHF Heart Protection Study (Parish et al., 2012), the best predictor of incident CVD in CKD patients (Shao et al., 2023), and predicted risk of CHD death in the MRFIT trial (Kuller et al., 2007). This is unlikely to be due to improved cholesterol efflux capacity (CEC): Shao and colleagues note that S-HDL-P is more efficient in promoting cholesterol efflux, and that M-HDL-P is associated with protection from vascular complications of type 1 diabetes without a corresponding difference in measured CEC (Shao et al., 2023). Other studies have shown inverse associations of L-HDL-P with incident CVD (Mora et al., 2009) and both M-HDL-P and L-HDL-P with incident coronary heart disease (Akinkuolie et al., 2014). Given the above, the stronger impact of this study’s focal interaction on larger HDL subfractions increases its clinical relevance beyond the previously recognized effect on HDL-C. Notably, women have higher HDL-C on average, as well as a larger mean particle size (Franczyk et al., 2023), linking to the observed sex dependence in the interaction effect. The potential for a common causal factor underlying these observations is an intriguing direction for further study.

We did not find evidence that the interaction explored here is specific to a certain intensity or domain of PA. Interaction effects using moderate versus vigorous PA were similar, and in UKB, PA estimates based on the IPAQ questionnaire (which additionally includes occupational PA) produced stronger interactions. PA has well-established positive associations with HDL cholesterol and function (Franczyk et al., 2023), and accumulating evidence points to a positive impact on larger HDL subclasses (Franczyk et al., 2023; Sarzynski et al., 2015), with directional consistency across exercise intensities (Slentz et al., 2007). These relationships are notably complex, however. While PA tends to increase L-HDL-P, it is consistently associated with modestly lower M-HDL-P (Sarzynski et al., 2015). Furthermore, baseline HDL characteristics may affect response to exercise; in individuals with prediabetes, M-HDL-P predicts the impact of exercise on HDL functionality, with implications for downstream cardiovascular risk (Sarzynski et al., 2018).

Meta-regression, motivated by the strength of the interaction in the all-female WGHS cohort, suggested that this interaction is more prominent in women. This may link to the HDL subfraction findings discussed above; women have higher HDL-C on average, as well as a larger mean particle size (Franczyk et al., 2023), which we confirmed in UKB. Though two-way interactions are already complex, these results indicate the importance of exploring three-way interactions for important potential effect modifiers such as sex. We note that the observation of female-specific effects allowed us to uncover interaction effects with larger magnitude in UKB than if we had assumed homogeneity of these interaction effects.

While interrogation of genetic mechanisms at the intergenic rs295849 was not the primary focus of this investigation, we present additional notes here. As originally reported by Kilpeläinen et al. (Kilpeläinen et al., 2019), the nearest gene is *LHX1*, encoding a transcription factor without clear relevance to lipoprotein metabolism. They suggest the nearby acetyl-CoA carboxylase (*ACACA*), encoding a key enzyme in fatty acid biosynthesis and metabolism pathways, as a biologically plausible effector gene. In support of this hypothesis, we find that a chromatin interaction was reported between one chromatin anchor harboring rs295849 and another at *ACACA* (Jin et al., 2013; Pan et al., 2021), suggesting that the observed statistical interaction may reflect co-regulation events involving *ACACA*. Despite rs295849 sitting outside a clear genic or enhancer region, its RegulomeDB score of 0.6 indicates potential regulatory activity, based partially on its binding by the transcription factor and spliceosome component RBM25 (Dong et al., 2023). RBM25 affects lipid metabolism; the overexpression of three rare RBM25 mutants in Huh-7 hepatocytes resulted in decreased LDL uptake (Zanoni et al., 2022). Furthermore, *RBM25* gene expression tends to be higher in females, providing a possible link to the sex-biased interaction effect observed here (Zhang 2021). Functional experiments exploring context-specific expression of potential effector genes (such as *ACACA*) and activity of RBM25 may help to clarify the molecular mechanisms at play.

This study’s major strength lies in the accumulation of evidence and refinement across multiple cohorts and data types, helping to expand and reinforce conclusions from each other and the originally reported interaction. Despite this fact and our attention to harmonization of PA and outcome variables, residual heterogeneity across cohorts remains a notable limitation. Questionnaire items informing calculated PA differed between the three cohorts here, which may have contributed to the differing shapes of the PA-HDL main effect relationship. As discussed, NMR platforms differed between UKB and the other studies, with imperfect ability to harmonize HDL subfractions across platforms using average particle diameters. Furthermore, in this study we did not investigate downstream links between HDL particles and risk for cardiovascular and other diseases; ongoing research into the effectiveness of HDL subfractions as predictive biomarkers may help to strengthen this link.

In conclusion, this study makes two key contributions. First, we provide substantially greater insight into a previously discovered gene-PA interaction, showing how rs295849 interacts with PA behaviors to impact lipoprotein profiles associated with cardiovascular risk. Second, we demonstrate that careful attention to phenotype refinement and harmonization, covariate adjustment, and relevant population subgroups can account for some sources of heterogeneity across studies and thus allow for improved GxE replication. This approach provides a template for future studies to characterize and strengthen GxE findings, which will be a critical ingredient in building a robust evidence base of gene-lifestyle interactions to inform precision medicine.

## Methods

### Women’s Genome Health Study

The WGHS is a prospective US-based cohort of healthy adult females with baseline age 45 years or older at enrollment in 1992-1994, and includes a majority subset with European ancestry verified by genetics that was used here and in the original analysis (Ridker et al., 2008,Kilpeläinen et al., 2019). The WGHS component of this work was conducted under approval from the Mass General Brigham IRB (protocol 2006P001259).

HDL-C was determined enzymatically from baseline plasma, and log transformed for analysis. Chemically-measured (rather than NMR-measured) HDL-C was used for analysis unless otherwise noted. Physical activity (PA) was ascertained by self-report questionnaire and dichotomized (a threshold of 225 MET-mins/wk moderate-to-vigorous PA) (refer to Kilpeläinen et al., 2019). We note that the original analysis coded physical activity as active (0) compared to inactive (1). Here, the coding in the WGHS is the opposite (active=1, inactive=0) so that the interpretation matches the rest of the current analysis. Genotype dosages for rs295849 were based on microarray genotyping followed by imputation to the TOPMed reference panel (August 26, 2019). Sensitivity models included one with five genetic principal components and another with genotype-covariate product terms for each covariate.

Covariates for the analysis included age at baseline, alcohol (four intake categories and other), smoking (current, former, never, or other), educational attainment (categorical as L.P.N. or L.V.N., 2-year R.N., 3-year R.N., Bachelor’s, Master’s, Doctoral, or other), income (categorical in thresholds $10,000, $20,000, $30,000, $40,000, $50,000, $100,000, and other), and a diet quality score based on the Alternative Healthy Eating Index. We note that this covariate set includes additional variables related to socioeconomic status (SES) and healthy lifestyle compared to those from the original meta-analysis.

45 metabolites measured using nuclear magnetic resonance (NMR) were available from the LipoScience/Labcorp Vantera^®^ Clinical Analyzer platform (subsequently referred to as “Labcorp” in this manuscript; using the LP4 algorithm) as previously described (Ahmad et al., 2018), reporting the concentrations of lipoprotein subfraction particles according to class (LDL, HDL, and triglyceride-rich lipoprotein particles [TRLP]) and physical diameter (Huffman et al., 2022).

### UK Biobank

UKB is a large prospective cohort with both deep phenotyping and molecular data, including genome-wide genotyping, on over 500,000 individuals of age 40-69 living throughout the UK between 2006-2010 (Sudlow et al., 2015). The UKB component of this work was conducted under a Not Human Subjects Research determination for UKB data analysis (NHSR-4298 at the Broad Institute of MIT and Harvard) and UK Biobank application 27892. We used a sample set including individuals of European ancestry (determined by the Pan-UKBB project (Pan-UKB team, 2020)) that had not withdrawn consent by the time of analysis. Additionally, we limited analysis to a subset of unrelated individuals (by including only those that were used for genetic principal components analysis during central genetic data preprocessing) and removed participants who were pregnant or had diabetes, coronary heart disease, liver cirrhosis, or cancer.

The outcome trait, HDL-C, was originally measured using enzyme immunoinhibition analysis in plasma samples and was log-transformed prior to analysis. Multiple PA exposures were derived from questionnaire responses. International physical activity questionnaire (IPAQ)-based PA estimates (MET-min/wk) were derived from touchscreen questions about the frequency and duration of moderate and vigorous intensity physical activity and walking (e.g., “Number of days/week of vigorous physical activity 10+ minutes”). Recent physical activity questionnaire (RPAQ)-based PA estimates (MET-min/wk) were derived from touchscreen questions about the frequency and duration of leisure-time physical activity in various specific categories (e.g., “Duration of strenuous sports”). Genotyping, imputation, and initial quality control for the UKB genetic dataset have been described previously (Bycroft et al., 2018). Variant rs295849 was genotyped directly and was retrieved from genetic data release version 3.

Covariates included genetically-determined sex, age, age^2^, a sex-by-age product term, 5 genetic principal components (calculated centrally using the entire UKB population), smoking (categorical: never, past, or current), alcohol intake (categorical: weekly frequency estimates), quantitative diet variables from a food frequency questionnaire (cooked vegetables, raw vegetables, fresh fruit, oily fish, non-oily fish, processed meat), a categorical diet variable (bread type), and a multiple deprivation index (one for each of England, Scotland, and Wales, with each variable containing a quantitative value for participants living in that country and zero otherwise). For variables coded as categorical, ambiguous categories such as “do not know” or “prefer not to answer” were left as non-missing to allow them to constitute an independent category for adjustment. All continuous variables, including blood biomarkers, PA variables, and continuous covariates, were winsorized at 5 standard deviations from the mean after all other preprocessing steps.

NMR metabolites were available from the Nightingale platform for 66,870 participants (having other necessary variables and passing described exclusion criteria) and were preprocessed using the *ukbnmr* R package (Ritchie et al., 2023), which includes imputation of zero values, log-transformation, adjustment for key batch variables such as shipment plate and time between sample preparation and measurement, and transformation back into absolute concentrations.

After preprocessing, 325 metabolites were available for analysis, covering a similar set of biological quantities (primarily lipoprotein lipid and particle concentrations) and including additional derived variables: 107 non-derived variables (e.g., M-HDL-P concentration), 61 composite variables (e.g., total lipids in medium HDL), 135 percentages (e.g., cholesterol as a percentage of total lipids in medium HDL), and 22 ratios (e.g., free cholesterol to cholesteryl esters in HDL).

### Multi-Ethnic Study of Atherosclerosis

The Multi-Ethnic Study of Atherosclerosis (MESA) is a study of subclinical cardiovascular disease and the risk factors that predict progression to clinically overt cardiovascular disease or progression of the subclinical disease (Bild, 2002). MESA consisted of a diverse, population-based sample of an initial 6,814 asymptomatic men and women aged 45-84, with the first examination (analyzed here) taking place between July 2000 and July 2002. 38 percent of the recruited participants were white, 28 percent African American, 22 percent Hispanic, and 12 percent Asian, predominantly of Chinese descent. The MESA component of this work was conducted under approval from the Mass General Brigham IRB (protocol 2017P000531).

As in WGHS, chemically-measured (using the cholesterol oxidase method [Roche Diagnostics, Indianapolis, Indiana]; rather than NMR-measured) HDL-C was used for analysis unless otherwise noted. The primary PA measure used in MESA was moderate and vigorous PA (MVPA), derived as previously described (Bertoni et al., 2009). Briefly, a series of questionnaire items (28 questions including household chores, lawn/yard/garden/farm, care of children/adults, transportation, walking (not at work), dancing and sport activities, conditioning activities, leisure activities, and occupational and volunteer activities) were used to generate weekly activity levels in three categories (light, moderate, and vigorous), with MVPA calculated as the simple sum of all moderate and vigorous activity. Genotypes in MESA came from whole-genome sequencing (WGS) data generated through the NHLBI TOPMed program (Freeze 9b data release). Details on WGS data generation and preprocessing are available at: https://topmed.nhlbi.nih.gov/topmed-whole-genome-sequencing-methods-freeze-9. Variant rs295849 was extracted for MESA participants from TOPMed-wide .gds files.

NMR-based lipoprotein subclasses were measured in 2012 using the LipoScience/Labcorp Vantera^®^ Clinical Analyzer platform (LP4 algorithm, matching that of WGHS) as previously described (Huffman et al., 2022). Missing covariate values were imputed to preserve sample size, using the median value for continuous variables, a missing indicator for categorical income, and “never” for smoking.

### Statistical modeling

For all cohorts, the primary interaction model used the following form:

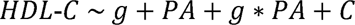

where g represents the genotype, PA represents physical activity, and C represents covariates. HDL-C was log-transformed in WGHS and MESA and treated as a continuous variable (log-transformation had minimal effect on model estimates). Covariates differed slightly between cohorts due to data availability, but aimed to adjust for age, age^2^, sex, healthy lifestyle, and socioeconomic status. The directed acyclic graph guiding these choices can be found in Supp. Fig. S4. PA main effect models omitted the genetic main effect and interaction product terms. When modeling NMR metabolites, the statistical model remained the same, with metabolite quantities replacing HDL-C as the outcome. Unless otherwise noted, data analysis was conducted using R version 4.2.2 (R Core Team, 2022).

Meta-regression of interaction summary statistics from Kilpelaïnen et al. was performed to test whether there was a systematic difference in gene-PA interaction effect estimates according to sex. Meta-regression used the *meta* package for R (*metareg* function), based on a generic inverse variance meta-analysis (*metagen* function) with variance estimation by restricted maximum likelihood and Wald-type confidence intervals. Sex proportions (between 0 [all male] and 1 [all female]) for each cohort-ancestry combination were not reported by Kilpeläinen et al., so were retrieved primarily from a prior study using these same cohorts (De Vries et al., 2019) or using literature-based reports for the few remaining cohorts otherwise. The primary meta-regression model included an intercept to avoid the strong assumption that the interaction effect is precisely zero in all-male cohorts. Sensitivity analyses included (1) exclusion of single-sex cohorts that will tend to exert high leverage on the regression fit, and (2) leave-one-out analyses that systematically excluded each study-population combination.

## Data and code availability

Code supporting the analyses described here can be found at https://github.com/kwesterman/gxpa-nmr. Access to WGHS data is restricted by the institutional review board, but analysis may be performed through collaboration; please contact Daniel Chasman. The UK Biobank data can be obtained through application at https://www.ukbiobank.ac.uk/. MESA data can be accessed through the TOPMed program via the NCBI Database of Genotypes and Phenotypes (dbGaP).

## Acknowledgments

This investigation was supported by two grants from the U.S. National Heart, Lung, and Blood Institute (NHLBI), the National Institutes of Health, R01HL118305 and R01HL156991. KEW was supported by K01DK133637. TOK was supported by the Novo Nordisk Foundation (NNF18CC0034900, NNF21SA0072102). ARB was supported by the Intramural Research Program of the National Human Genome Research Institute of the National Institutes of Health through the Center for Research on Genomics and Global Health (CRGGH). SM was supported by HL160799, HL117861, and K24 HL136852.

The WGHS is supported by the National Heart, Lung, and Blood Institute (HL043851 and HL080467) and the National Cancer Institute (CA047988 and UM1CA182913), with funding for genotyping provided by Amgen and funding for NMR assays by the American Heart Association.

Whole genome sequencing (WGS) for the Trans-Omics in Precision Medicine (TOPMed) program was supported by the National Heart, Lung and Blood Institute (NHLBI). WGS for “NHLBI TOPMed: Multi-Ethnic Study of Atherosclerosis (MESA)” (phs001416.v3.p1) was performed at the Broad Institute of MIT and Harvard (3U54HG003067-13S1). Centralized read mapping and genotype calling, along with variant quality metrics and filtering were provided by the TOPMed Informatics Research Center (3R01HL-117626-02S1). Phenotype harmonization, data management, sample-identity QC, and general study coordination, were provided by the TOPMed Data Coordinating Center (3R01HL-120393-02S1), and TOPMed MESA Multi-Omics (HHSN2682015000031/HSN26800004). The MESA projects are conducted and supported by the National Heart, Lung, and Blood Institute (NHLBI) in collaboration with MESA investigators. Support for the Multi-Ethnic Study of Atherosclerosis (MESA) projects are conducted and supported by the National Heart, Lung, and Blood Institute (NHLBI) in collaboration with MESA investigators. Support for MESA is provided by contracts 75N92020D00001, HHSN268201500003I, N01-HC-95159, 75N92020D00005, N01-HC-95160, 75N92020D00002, N01-HC-95161, 75N92020D00003, N01-HC-95162, 75N92020D00006, N01-HC-95163, 75N92020D00004, N01-HC-95164, 75N92020D00007, N01-HC-95165, N01-HC-95166, N01-HC-95167, N01-HC-95168, N01-HC-95169, UL1-TR-000040, UL1-TR-001079, UL1-TR-001420, UL1TR001881, DK063491, and R01HL105756. The authors thank the other investigators, the staff, and the participants of the MESA study for their valuable contributions. A full list of participating MESA investigators and institutes can be found at http://www.mesa-nhlbi.org.

## Supplementary Figures

**Supplementary Figure S1:**
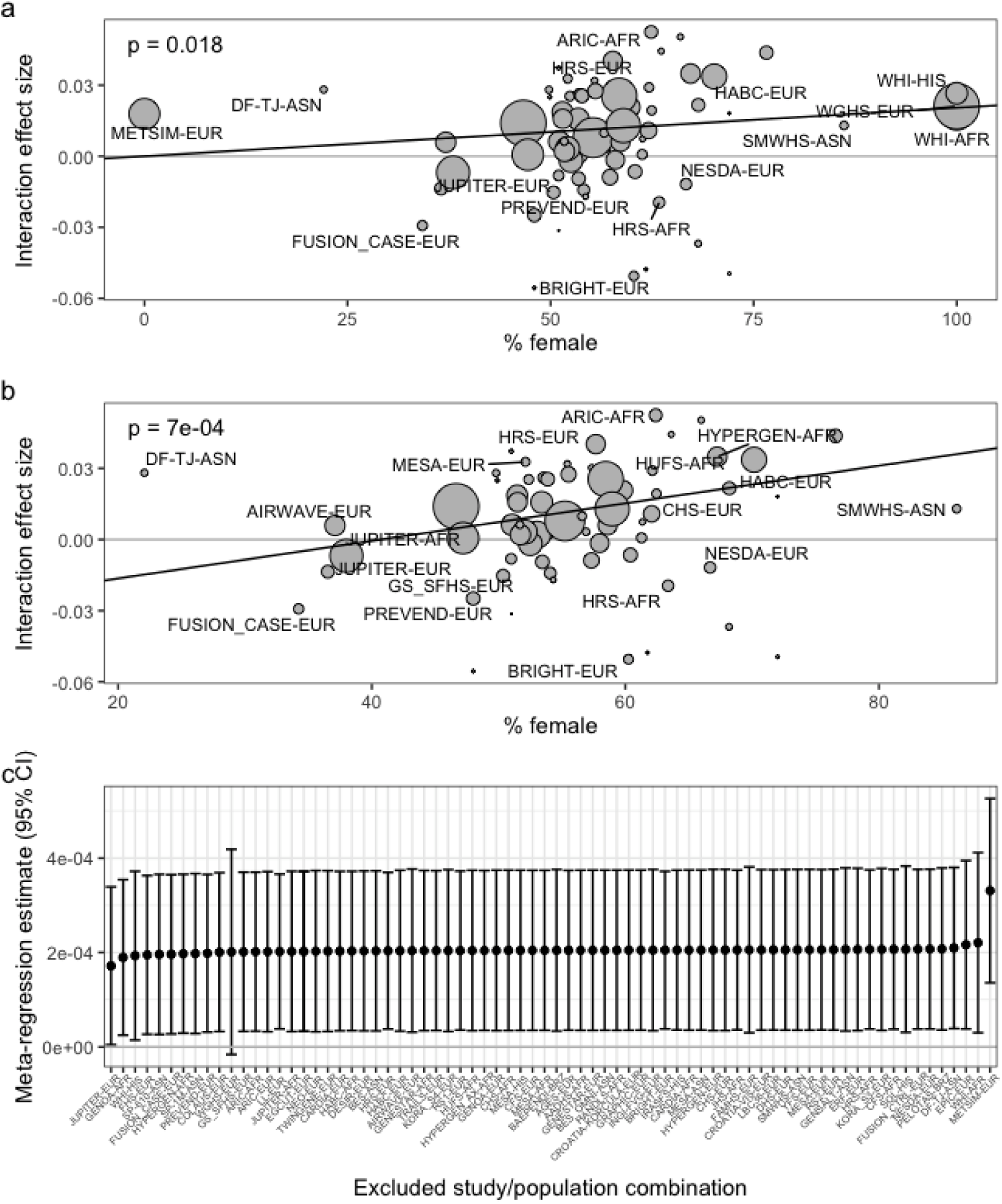
Bubble plot describing the meta-regression of interaction summary statistics from Kilpeläinen et al. a) Interaction effect sizes from each cohort are plotted against the corresponding sex proportions for the primary meta-regression. Bubble size is proportional to the inverse variance of the interaction effect estimate. Study labels are shown for studies with N > 1,000. b) As in (a), but excluding all single-sex studies. c) Leave-one-out sensitivity analysis plots show meta-regression estimates and confidence intervals after excluding each study-population combination.

**Supplementary Figure S2:**
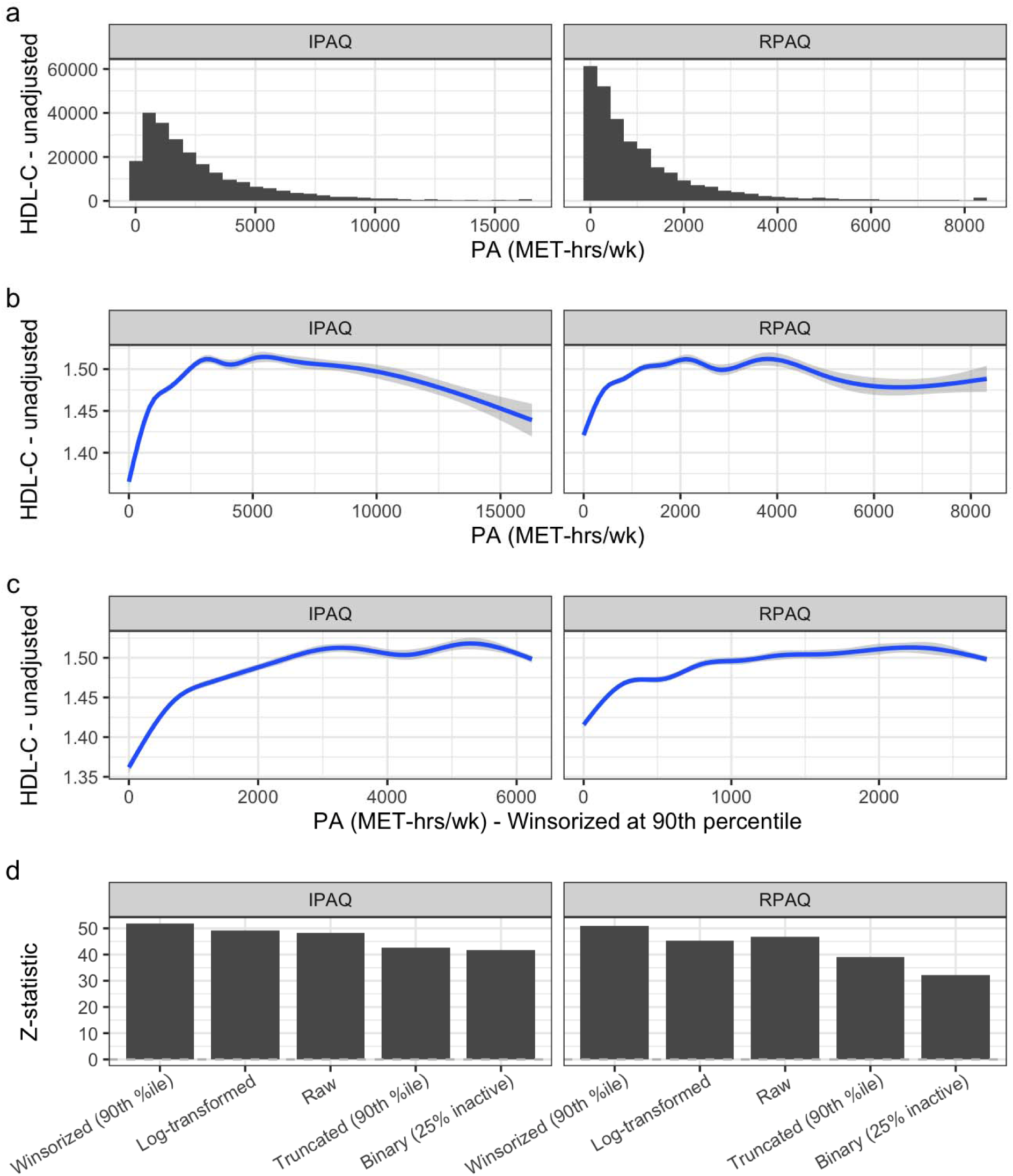
PA-HDL-C main effects in UKB. a) Histogram of PA measured using questions based on the IPAQ (left) or RPAQ (right) questionnaires. b) Shrunken cubic spline fits of the relationship between PA and unadjusted mean HDL-C, with shadows corresponding to 95% CIs. c) As in (b), but using PA distributions winsorized at the 90^th^ percentile. d) Adjusted main effect z-statistics according to PA transformation.

**Supplementary Figure S3:**
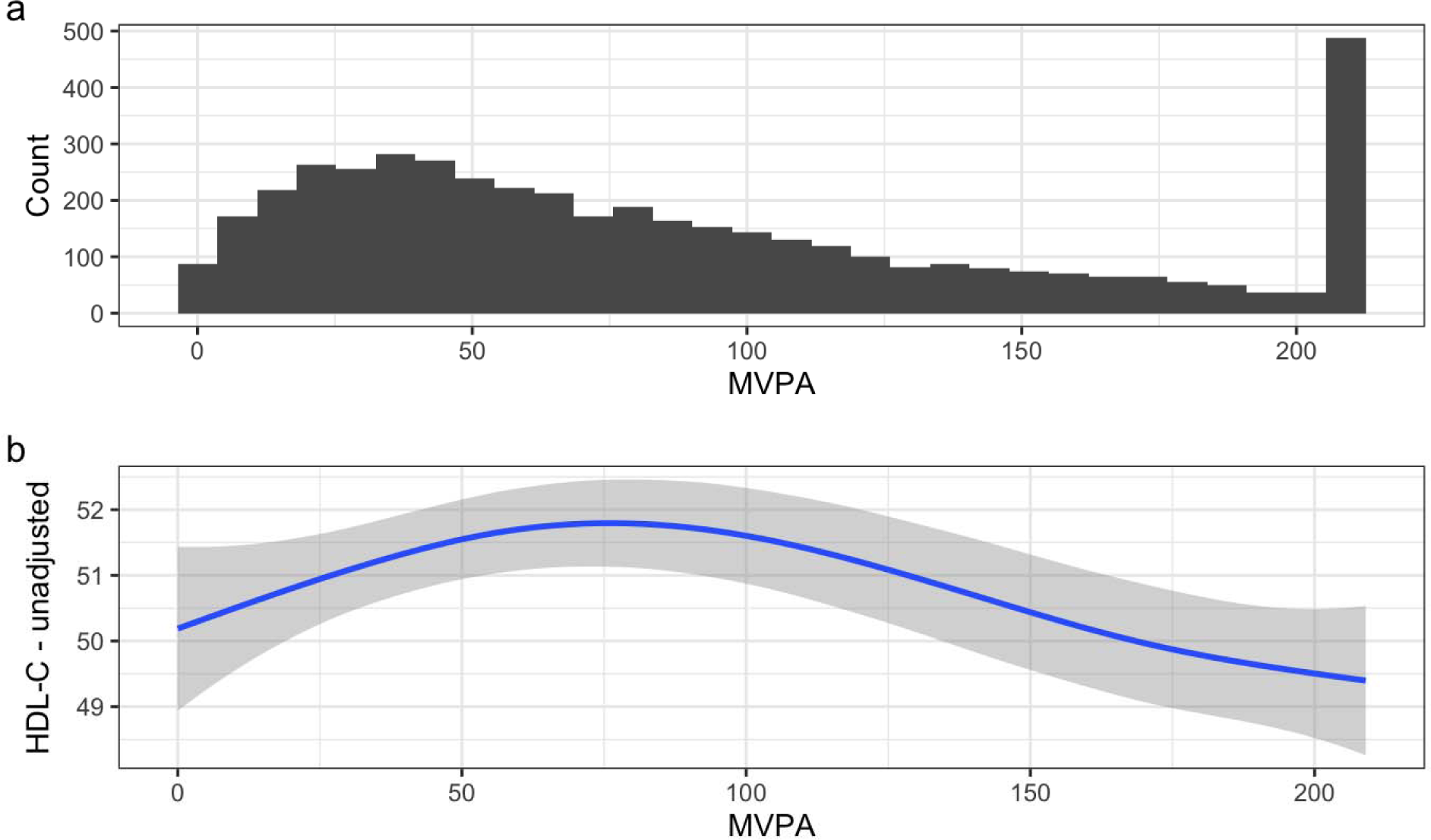
PA-HDL-C main effects in MESA. a) Histogram of questionnaire-based moderate-to-vigorous PA estimates. b) Shrunken cubic spline fit of the relationship between PA and unadjusted mean HDL-C, with shadows corresponding to 95% CIs.

**Supplementary Figure S4:**
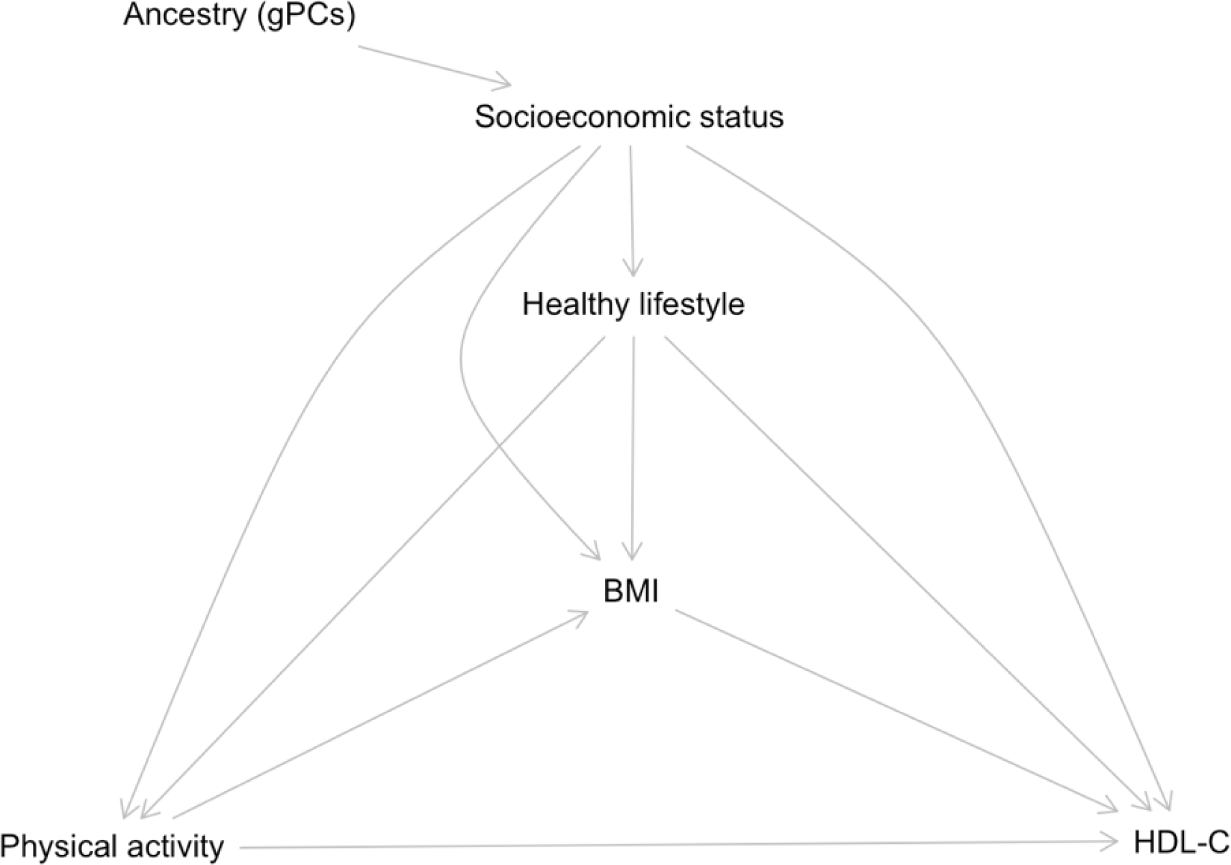
Directed acyclic graph informing covariate selection based on PA-HDL-C main effect.

## Supplementary Tables

See accompanying Excel document.

